# Knowledge of COVID-19 prevention in Eastern Ethiopia

**DOI:** 10.1101/2023.06.28.23291972

**Authors:** Merga Dheresa, Zachary J. Madewell, Jonathan A. Muir, Tamirat Getachew, Gamachis Daraje, Gezahegn Mengesha, Cynthia G. Whitney, Nega Assefa, Solveig A. Cunningham

## Abstract

**Objectives:** As of May 2023, over 500,000 COVID-19 cases and over 7,500 deaths have been reported in Ethiopia. Understanding community members’ knowledge and perception of SARS-CoV-2 prevention is essential for directing public health interventions to reduce transmission and improve vaccination coverage. Here, we aimed to describe factors associated with knowledge of COVID-19 prevention among community residents in Eastern Ethiopia.

**Methods:** We conducted a cross-sectional survey among a random sample of 880 participants in a Health and Demographic Surveillance System in the Harari Region, Ethiopia, from August to September 2021. Principal components analysis was used to create a score representing knowledge of COVID-19 prevention. Quasi-Poisson regression was used to examine associations between demographic characteristics and knowledge of COVID-19 prevention. Our survey also included information regarding knowledge of community or government measures to prevent COVID-19, healthcare services for children under five, and healthcare services for pregnant women.

**Results:** The most cited individual measures to reduce the risk of contracting COVID-19 were washing hands with soap (91.5%) and wearing a facemask (89.2%), whereas least mentioned were avoiding domestic and international travel (22.2%) and wearing medical gloves (20.3%). The most recognized community or government measures to prevent SARS-CoV-2 transmission were closure of schools and universities (77.0%), advice to avoid gatherings (75.2%), and advice to stay home (62.3%). Adjusted analyses demonstrated that knowledge of COVID-19 prevention was higher among participants from rural areas than urban areas, those aged ≥65 years (<25 years as reference), with secondary education (no formal education as reference), with monthly income of ≥2,001 Birr (0-1,200 as reference), and were farmers or domestic/subsistence workers or government employees (unemployed as reference). Knowledge was lower among households with ≥5 household members (1-2 as reference). Of households with children under five and pregnant women, 9.4% and 12.3% missed at least one medical care visit since mid-March 2020 consequent to the pandemic, respectively.

**Conclusions:** Public health interventions to reduce infectious disease transmission depend on perceptions of risk and knowledge. The survey found that most adults had good knowledge of methods for reducing risks of COVID-19, although knowledge differed between groups. A substantial number of respondents reported missing important healthcare visits. Understanding these factors may help Ethiopian authorities plan effective health education programs to control community and household transmission of SARS-CoV-2.

## Introduction

Coronavirus disease 2019 (COVID-19) was first reported in Ethiopia in March 2020. Since then, over 500,000 laboratory-confirmed cases and over 7,500 deaths have been reported in Ethiopia through May 2023, accounting for 5.2% of total COVID-19 cases and 4.3% of COVID-19 deaths reported in Africa.^1^ However, COVID-19 tests were largely unavailable in Ethiopia throughout the pandemic, and as of March 2022, only three million COVID-19 tests had been performed among Ethiopia’s 120 million population, implying that the actual number of cases is likely much higher.^2^

Knowledge, attitude, and practices (KAP) studies of COVID-19 in sub-Saharan Africa have demonstrated positive associations between knowledge of COVID-19 symptoms, transmission, and prevention, and implementation of public health measures such as hand hygiene, social distancing, and facemask use.^3, 4^ Knowledge of COVID-19 symptoms, transmission, and prevention has also been positively associated with vaccination coverage,^5–7^ which is one of the most effective strategies for protecting individuals against COVID-19 hospitalization and death.^8, 9^ COVID-19 vaccines were introduced in Ethiopia in March 2021, initially prioritizing healthcare workers, older adults, and individuals with chronic diseases.^10^ The vaccination campaign was expanded in November 2021 to include all individuals aged 12 years or older.^11^ By May 2023, 32.6% of Ethiopia’s population had completed a primary COVID-19 vaccine series of BBIBP-CorV (Sinopharm, Beijing CNBG) (two doses), BBV152 (Bharat Biotech’s Covaxin) (two doses), Ad26.COV2.S (Janssen) (one dose), or ChAdOx1-S (Covishield) (two doses), but only 2.5% had received a booster dose.^1^

How much the community knows about COVID-19 prevention is unclear for eastern Ethiopia. One community-based cross-sectional survey of adults in the Harari Region in February 2021 found low perceived risk of COVID-19—35.6% of participants indicated they would seek healthcare if they developed COVID-19 symptoms,^12^ although these data were collected before the emergence of more transmissible SARS-CoV-2 variants.^13, 14^ The authors found that perception of risk toward COVID-19 was greater among respondents from rural areas than urban areas, but they did not evaluate factors associated with knowledge of COVID-19 prevention among urban and rural residents, knowledge of community or government measures to prevent COVID-19, nor healthcare access among children under five and pregnant women during the pandemic. We conducted a community-based cross-sectional study to evaluate factors associated with knowledge of COVID-19 prevention in urban and rural settings of Eastern Ethiopia. Information resulting from this study will inform prioritizing health education programs regarding public health measures to prevent transmission of infectious disease among community residents.

## Methods

### Study setting

The study was conducted in a predominantly rural area in the Kersa District and an urban area in the Harari People’s National Regional State in Eastern Ethiopia. Both areas have been monitored through a Health and Demographic Surveillance System since 2012, with demographic and health-related information regularly collected.^15, 16^ The rural area comprises 24 kebeles (neighborhoods), covering 353 km^2^ and with a population of 135,754 in 25,653 households (Figure 1). The urban area comprises 12 kebeles, covering 25.4 km^2^ and with a population of 55,773 in 14,768 households.

**Figure 1.**
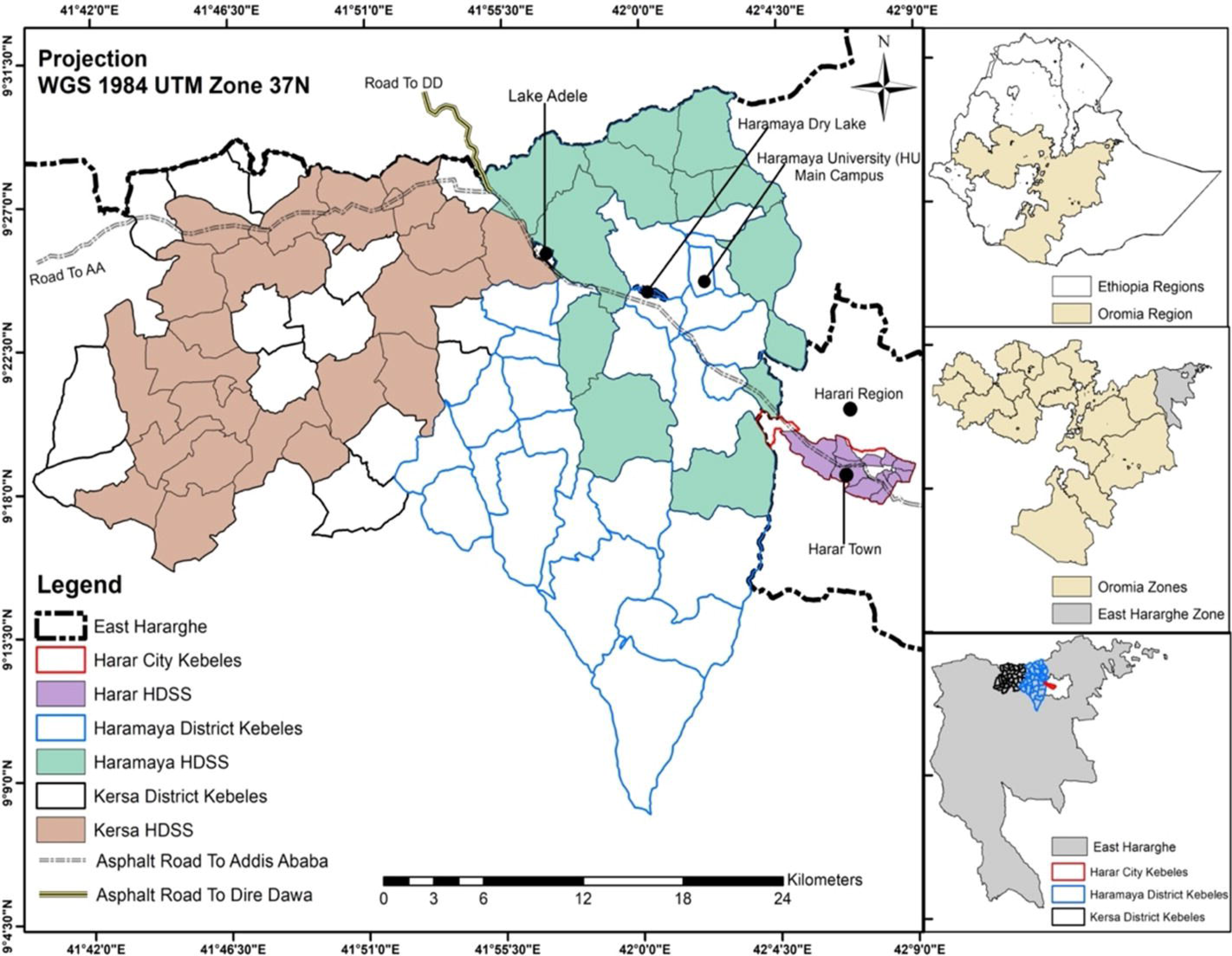
Geospatial Distribution of the Harar and Kersa Health and Demographic Surveillance Systems (HDSS) within East Hararghe, Oromia, Ethiopia. The smaller map panels on the right identify the location of the HDSS catchment areas within the East Haraghe Zone of the Oromia region in Ethiopia. The HDSS catchment in Haramaya (depicted in green) was in development during the data collection period, so households from this catchment were not included in this study.

### Study design

This study is part of a larger examination within the Child Health and Mortality Prevention Surveillance (CHAMPS) network to understand the consequences of COVID-19 lockdowns for child health and mortality.^15–17^ A survey instrument was developed to collect information about households’ experiences during the COVID-19 lockdown, including their knowledge of personal and community or government measures to prevent COVID-19, access to healthcare services for under-five children, and healthcare services for pregnant women.

Households were selected through simple random sampling from a sample frame of all households in the HDSS to achieve a sample size of 880 participants: 440 from four kebeles in the rural catchment area (Kersa) and 440 from four kebeles in the urban catchment area (Harar) (Figure 1). Sample weights were not applied. The sample size was specified *a priori* to detect prevalence of changes in accessing healthcare, with specifications of 50% of the population experiencing changes, 95% confidence interval (CI), precision of 0.05, and non-response adjustment of 10%. Data collectors were selected from the fieldwork teams of HDSS enumerators, who were already trained and working in the HDSS. Data collection occurred between August and September 2021 and was carried out through tablet-based, in-person interviews with adult household members. All of the 880 sampled households consented and participated in the survey. Data from the questionnaire were linked with data from the most recent completed HDSS round (collected from January to May 2020) to incorporate additional demographic data about the sampled household, specifically: number of children under 5 years of age, number of adults over age 60 in the household, household assets and residence construction materials. Data quality assurance and cleaning followed standard procedures for the HDSS,^18, 19^ and inconsistent or missing data were flagged for data collectors to correct. Field supervisors and the field coordinator selected a random sub-sample of questionnaires for re-visits to validate the recorded information. The data used are accessible at UNC Dataverse.^20^ Implementation of the module was approved by the Institutional Health Research Ethics Review Committee (IHRERC) with approval reference number Ref.No.IHRERC/127/2021.

## Measures

Interviewers asked which measures can be adopted to reduce the risk of contracting COVID-19 without providing options and recorded participants’ responses. Responses were subsequently categorized as: handwashing, sanitizer use, avoiding handshaking/physical greeting, mask use, medical gloves use, avoiding travel, avoiding going out, avoiding crowded places, two-meter social distancing, and other.

This survey included a closed-ended question asking participants which steps the community or government had taken to prevent the spread of SARS-CoV-2. Interviewers read each of the following options aloud and recorded participants’ responses: advising citizens to stay home, avoiding gatherings, restricting travel within country/area, restricting international travel, closing of schools and universities, imposing curfew or lockdown, closing of non-essential businesses, conducting sensitization or public awareness campaigns, establishing isolation centers, and disinfecting of public places.

Individual characteristics included: sex (female, male), age group (<25 years, 25-44 years, 45-64 years, ≥65 years), residence (urban, rural), ethnicity (Oromo, Amhara, other), religion (Christian, Muslim, other), marital status (married, separated/divorced, widowed, single), education (no formal education, primary, secondary, college), occupation (unemployed, farmer or domestic/subsistence worker, government employee, private employee, farmer, other), and health insurance (yes, no). Household variables were: household size (1-2, 3-4, 5-6, 7-8, ≥9), children under five years (yes, no), adults over 60 years (yes, no), pregnant women (yes, no), household member tested positive for SARS-CoV-2 (yes, no), monthly income (≥4,600, 3,001-4,600, 2,001-3,000, 1,201-2,000, 0-1,200 Birr, which is roughly equivalent in U.S. dollars to ≥$127, $83-127, $55-83, $33-55, and $0-33 ($1 USD ≈ 36.4 Birr at the time of this survey in August-September 2021), and television ownership (yes, no).

For participants with children under five, our survey asked whether those children had attended any healthcare visits between mid-March 2020 (yes, no) and when the survey was conducted in August-September 2021, and whether they needed medical care or a clinic visit but could not do so since mid-March 2020 (yes, no). We recorded the kind of medical care the child received or needed but did not receive (routine follow-up visits, routine vaccinations, clinic visits for any illness, services for malnutrition), the number of missed medical care or clinic visits, and reasons for not receiving healthcare (clinic closed, out of vaccines or medications, did not get transportation, lockdown, afraid to go).

For households with pregnant women between mid-March 2020 and August-September 2021, we asked whether they had attended any pregnancy-related healthcare since mid-March (yes, no) and whether they needed medical care during pregnancy but did not receive it (yes, no). We also recorded the kind of healthcare received or needed but not received (routine antenatal care visits, pregnancy-related complication or concern, delivery, Caesarean section, illness not related to pregnancy, medications, routine postanal care visit, postnatal concern or complications), the number of missed medical care or clinic visits, and reasons for not receiving healthcare (out of medications, lockdown, afraid to go).

### Statistical Analysis

We presented frequency distributions of individual characteristics (age, sex, residence, ethnicity, religion, marital status, education, occupation, has health insurance), household characteristics (household size, children under five, adults over 60, pregnant women, income, television, household member tested positive for SARS-CoV-2), child healthcare access (had children under five attend healthcare visits, needed medical care but could not do so, number of visits missed, reasons for not receiving healthcare), and pregnancy healthcare access (attended pregnancy-related healthcare, needed medical care but could not get it, number of visits missed, reasons for not receiving healthcare). We reported frequencies and 95% confidence intervals (95% CI) for knowledge of individual and community or government measures to prevent COVID-19. Given the differences in COVID-19 knowledge reported by urban and rural residents,^12^ we presented all results for the total population and stratified by urban/rural residence. Pearson Chi-square tests were used to evaluate associations between demographic characteristics and urban or rural residence.

In accord with other KAP studies,^21–25^ we used principal components analysis (PCA) to create a score for assessing the level of knowledge of COVID-19 prevention among all prevention variables, excluding other and gloves use, which may offer limited protection against SARS-CoV-2 transmission among community members.^26^ The overall Kaiser-Meyer-Olkin index of sampling adequacy was 0.89, indicating that the sample size and data were sufficient for PCA. Scores of one were assigned to an option if a respondent mentioned it and zero if they did not mention it. The resulting compound factor accounted for 46% of the variability in the data and included all variables (Table S1). These variables were then weighted against their eigenvector coefficients. Knowledge of prevention scores ranged from 0 to 4.5, with higher scores representing greater knowledge.

Quasi-Poisson regression was used to evaluate unadjusted and adjusted associations between characteristics (age group, sex, residence, ethnicity, religion, marital status, education, occupation, monthly income, health insurance, household size, children under five, adults over 60, pregnant women, television) and PCA-derived knowledge of prevention index for 1) all participants and 2) stratified by urban/rural residence. Variables were selected for inclusion in the final adjusted regression models if their unadjusted associations with knowledge of COVID-19 prevention had theoretical justification. Other studies have demonstrated significant associations between COVID-19 knowledge and age;^25, 27, 28^ sex;^25, 28^ ethnicity;^25^ religion;^25, 29^ marital status;^25, 28, 30^ education;^25, 31^ occupation;^25, 29^ income;^25^ health insurance;^32^ household size;^33, 34^ older age;^25^ and television as a source of COVID-19 information.^25^ Therefore, all variables from unadjusted analyses were included in the final model. Values of *p*<0.05 were considered statistically significant. We used the “car” package in R to derive generalized variance inflation factors (GVIF(1/(2*df)) for all independent variables in the adjusted model, where df is the degrees of freedom associated with each term.^35^ Generalized variance inflation factors were all <1.7 so there was no evidence of collinearity among independent variables (Table S2). We evaluated residual deviance as a goodness-of-fit test for the overall model. The residual difference was 381.3 (*p*=1.0), so we concluded that the model fit was reasonable. All analyses were done in R software, version 4.3.0 (R Foundation for Statistical Computing, Vienna, Austria).

## Results

Of all 880 participants, 81.9% were female, 65.7% identified as Oromo ethnicity, 69.1% were Muslim, 67.0% were married and 18.8% widowed, 50.2% had completed at least primary education, and 42.1% had health insurance. Regarding households, 43.8% had children under the age of five, 26.8% had adults over the age of 60, and 13.0% had pregnant women (Table 1). The median age of urban participants (45 years; interquartile range [IQR]: 35–60) was higher than rural participants (39 years; IQR: 30–48) (*p*<0.001). Two hundred sixty-six participants (30.2%) had family members who were ever tested for COVID-19, of whom 24 (9.0%) had at least one family member test positive. Compared to urban residents, a larger proportion of rural residents were Oromo (92.5% vs. 38.9%), Muslim (90.5% vs. 47.7%), married (79.1% vs. 55.0%), had no formal education (72.3% vs. 27.3%), and worked as a farmer or in a domestic/subsistence occupation (53.0% vs. 4.5%) (*p-*values≤0.035). The median household size in rural Kersa (6, IQR: 4–7) was larger than urban Harar (4, IQR: 3–5) (*p*<0.001). Furthermore, compared to urban households, a greater proportion of rural households had children under the age of five and pregnant women, whereas a smaller proportion had adults over 60 and SARS-CoV-2 positive household members (*p-*values≤0.006). More urban residents (n=384, 87.3%) owned a television than rural residents (n=85, 19.3%) (*p*<0.001). Median monthly income in urban Harar (3,000 Birr, IQR: 1,500, 50,00) was higher than rural Kersa (2,000 Birr, IQR: 1,300, 3,500) (*p*<0.001).

**Table 1.**
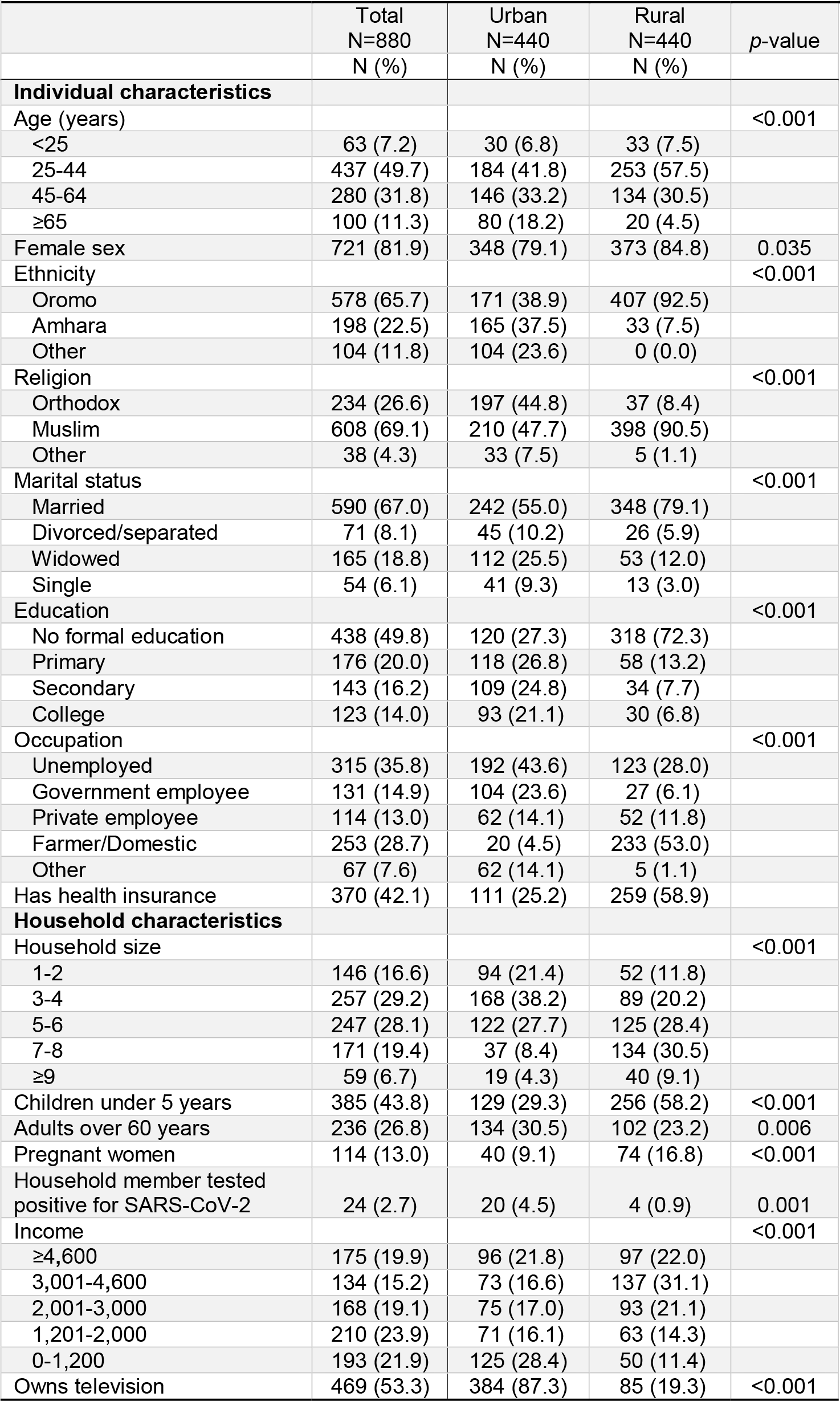
Descriptive statistics of individuals who participated in COVID-19 cross-sectional survey, Ethiopia, August – September 2021, (N=880)

All but six of the participants (99.3%, 874/880) had heard of COVID-19. The most frequently mentioned community or government measures to prevent SARS-CoV-2 transmission among all participants were closure of schools and universities (77.0%), advice to avoid gatherings (75.2%), and advice to stay home (62.3%), whereas least mentioned were restricted international travel (42.5%), curfew/lockdown (43.0%), and establishment of isolation centers (43.5%) (Figure S1). A greater proportion of urban participants than rural participants cited school closures, advice to stay home, and sensitization/public awareness as community or government prevention measures (*p-*values<0.001) (Figure 2).

**Figure 2.**
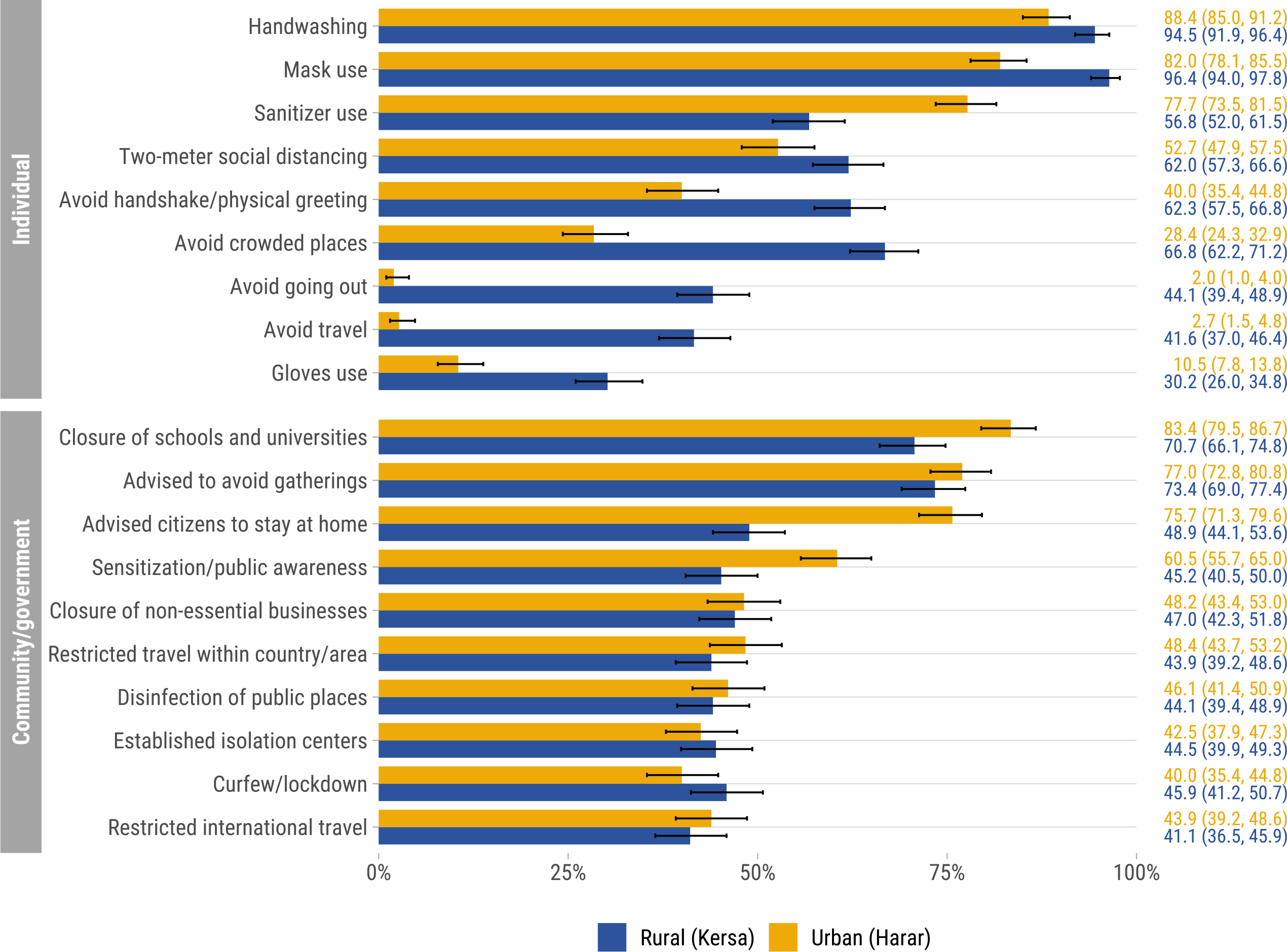
Knowledge of individual and community/government measures to prevent COVID-19 stratified by urban (Harar) and rural (Kersa) residence, Ethiopia, August – September 2021 (N=880). Error bars represent 95% confidence intervals.

Among all participants, the most commonly mentioned individual measures to reduce the risk of contracting COVID-19 were washing hands with soap (91.5%), wearing a facemask (89.2%), and using hand sanitizer (67.2%), whereas least mentioned were wearing medical gloves (20.3%), avoiding domestic and international travel (22.2%), and avoiding going out unless necessary (23.1%) (Figure S1). A greater proportion of rural than urban residents mentioned avoiding going out (44.1% vs. 2.0%), avoiding travel (41.6% vs. 2.7%), and avoiding crowded places (66.8% vs. 28.4%) (*p-*values<0.001) (Figure 2). Moreover, a greater proportion of rural than urban participants cited handwashing, mask use, two-meter social distancing, avoiding handshakes or physical greetings, and wearing medical gloves, whereas fewer mentioned using hand sanitizers (*p-*values≤0.001).

Unadjusted analyses between demographic characteristics and PCA-derived knowledge of prevention overall for all participants and stratified by urban/rural residence are shown in Figures 3 and S2, respectively. Adjusting for all other variables in the model (Figure 3), the knowledge of COVID-19 prevention score was -0.77 (95% CI: -0.86, -0.69) lower for participants from urban areas than those from rural areas. Monthly income of ≥4,600, 3,001-4,600, or 2,001-3,000 Birr (with 0-1,200 Birr as reference) had the strongest positive association with knowledge of COVID-19 prevention (β: 0.13 ∼ 0.67). COVID-19 prevention knowledge was also higher among participants who were ≥65 years (β: 0.38, 95% CI: 0.19, 0.57) with <25 years as reference, were farmers or worked in a domestic/subsistence occupation (β: 0.24, 95% CI: 0.15, 0.33) or were government employees (β: 0.14, 95% CI: 0.01, 0.27) with unemployed as reference, were Christian (β: 0.16, 95% CI: 0.05, 0.27) with Muslim as reference, were single (β: 0.18, 95% CI: 0.02, 0.33) with married as reference, and had secondary education (β: 0.17, 95% CI: 0.06, 0.29) with no formal education as reference. Conversely, knowledge of COVID-19 prevention was lower among households with 5-6, 7-8, or ≥9 household members (β: -0.29 ∼ -0.18). The results stratified by residence generally aligned with the overall findings, except for the positive association of older age and the inverse association of household size with COVID-19 knowledge, which were only significant for participants residing in rural areas (*p*-values≤0.001) (Figure S3). Furthermore, secondary education and Christian religion were only positively associated with COVID-19 knowledge for participants residing in urban areas (*p*-values≤0.029).

**Figure 3.**
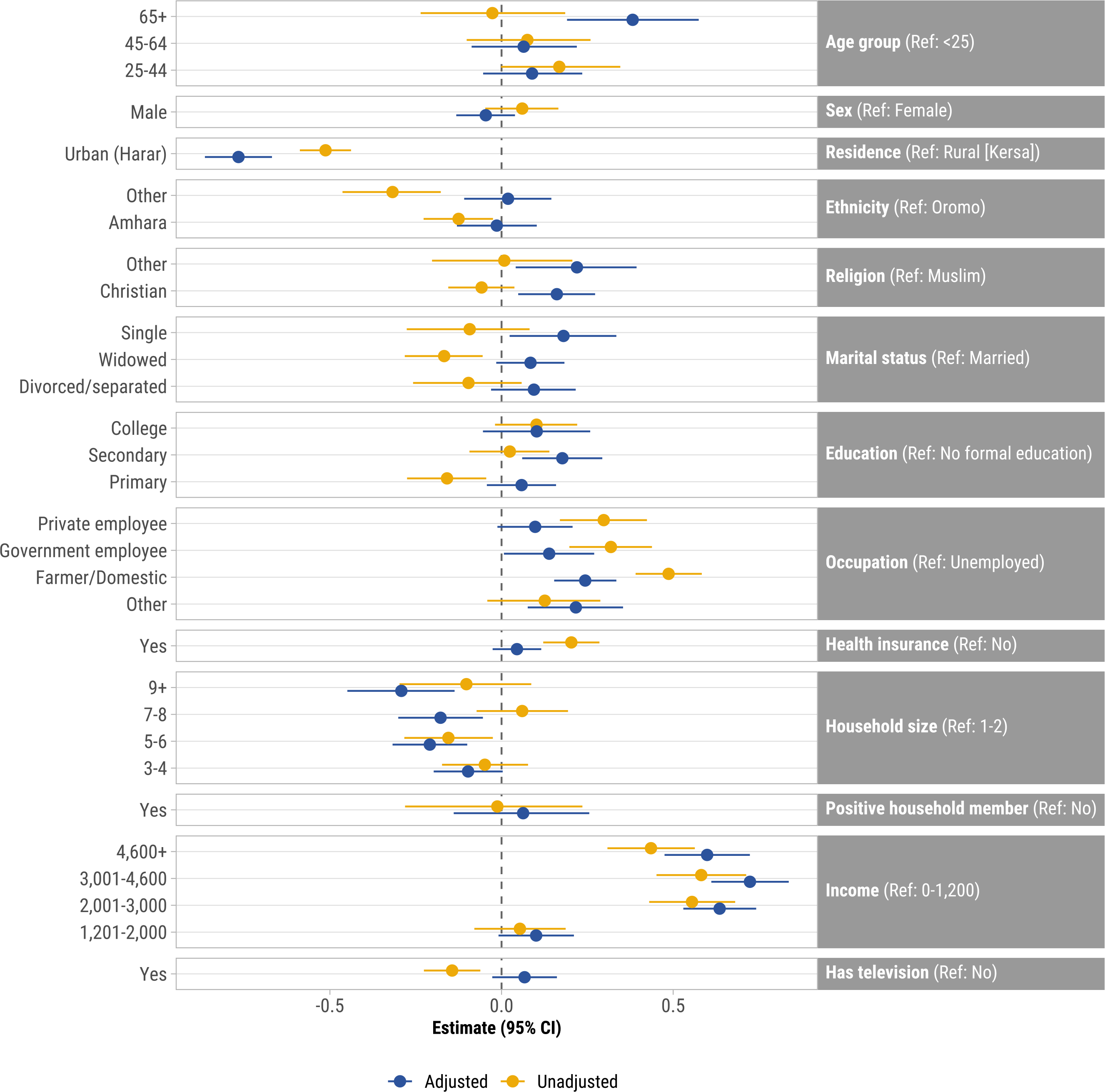
Unadjusted and adjusted associations between demographic characteristics and knowledge of COVID-19 prevention index, all participants (N=880), Ethiopia, August – September 2021. Points represent β coefficients and error bars represent 95% confidence intervals. Adjusted for all other variables in the model. Knowledge of prevention was derived from principal components analysis and includes: handwashing, sanitizer use, mask wearing, avoiding handshaking/physical greeting, avoiding travel, avoiding going out, avoiding crowded places, and two-meter social distancing.

Over half (59.0%, 227/385) of households with children under five attended child healthcare services between mid-March 2020 and August-September 2021, for reasons including routine or follow-up visits (41.9%, 95/227), illnesses (41.4%, 94/227), and vaccinations (33.9%, 77/227) (Table 2). However, thirty-six (9.4%) households needed medical care since mid-March 2020 but did not receive it. The missed care included vaccinations (63.9%, 23/36) and clinic visits for any illness (41.7%, 15/36). Reasons for not receiving healthcare services were fear of going to the clinic (61.1%, 22/36), lockdown restrictions (22.2%, 8/36), and shortage of vaccines or medications (19.4%, 7/385); only three (0.8%) did not receive care because the clinic was closed. The median number of visits missed was 2 (IQR: 1–3).

**Table 2.** Child healthcare access, Ethiopia, August – September 2021 (N=385)

|  | N | % |
| --- | --- | --- |
| Had children under 5 attend any healthcare visits since mid-March 2020 | 227 | 59.0 |
| Routine follow-up visit | 95 | 24.7 |
| Childhood vaccinations | 77 | 20.0 |
| Clinic visits for any illness | 94 | 24.4 |
| Services for malnutrition | 8 | 2.1 |
| Needed medical care/clinic visit but could not do so since mid-March 2020 | 36 | 9.4 |
| Routine follow-up visit | 7 | 1.8 |
| Routine vaccinations | 23 | 6.0 |
| Clinic visits for any illness | 15 | 3.9 |
| Services for malnutrition | 2 | 0.5 |
| Number of medical care/clinic visits missed |  |  |
| 1 | 9 | 2.3 |
| 2 | 11 | 2.9 |
| ≥3 | 16 | 4.2 |
| Reasons for not receiving healthcare<sup>a</sup> |  |  |
| Clinic closed | 3 | 0.8 |
| Out of vaccines or medications | 7 | 1.8 |
| Did not get transportation | 5 | 1.3 |
| Lockdown | 8 | 2.1 |
| Afraid to go | 22 | 5.7 |
| No response | 1 | 0.3 |

<sup>a</sup> Participants could provide multiple reasons

Almost three-quarters (73.7%, 84/114) of households with pregnant women had a pregnancy-related healthcare visit between mid-March 2020 and August-September 2021, for reasons including pregnancy-related complications or concerns (75.0%, 63/84), routine antenatal care visits (47.6%, 40/84), and delivery (32.1%, 27/84) (Table 3). Fourteen (12.3%) pregnant women required medical care during pregnancy since mid-March 2020 but did not receive it; the needed care included pregnancy-related complications (42.9%, 6/14) and routine antenatal care visits (28.6%, 4/14). The median number of visits missed was 1 (IQR: 1–2).

**Table 3.**
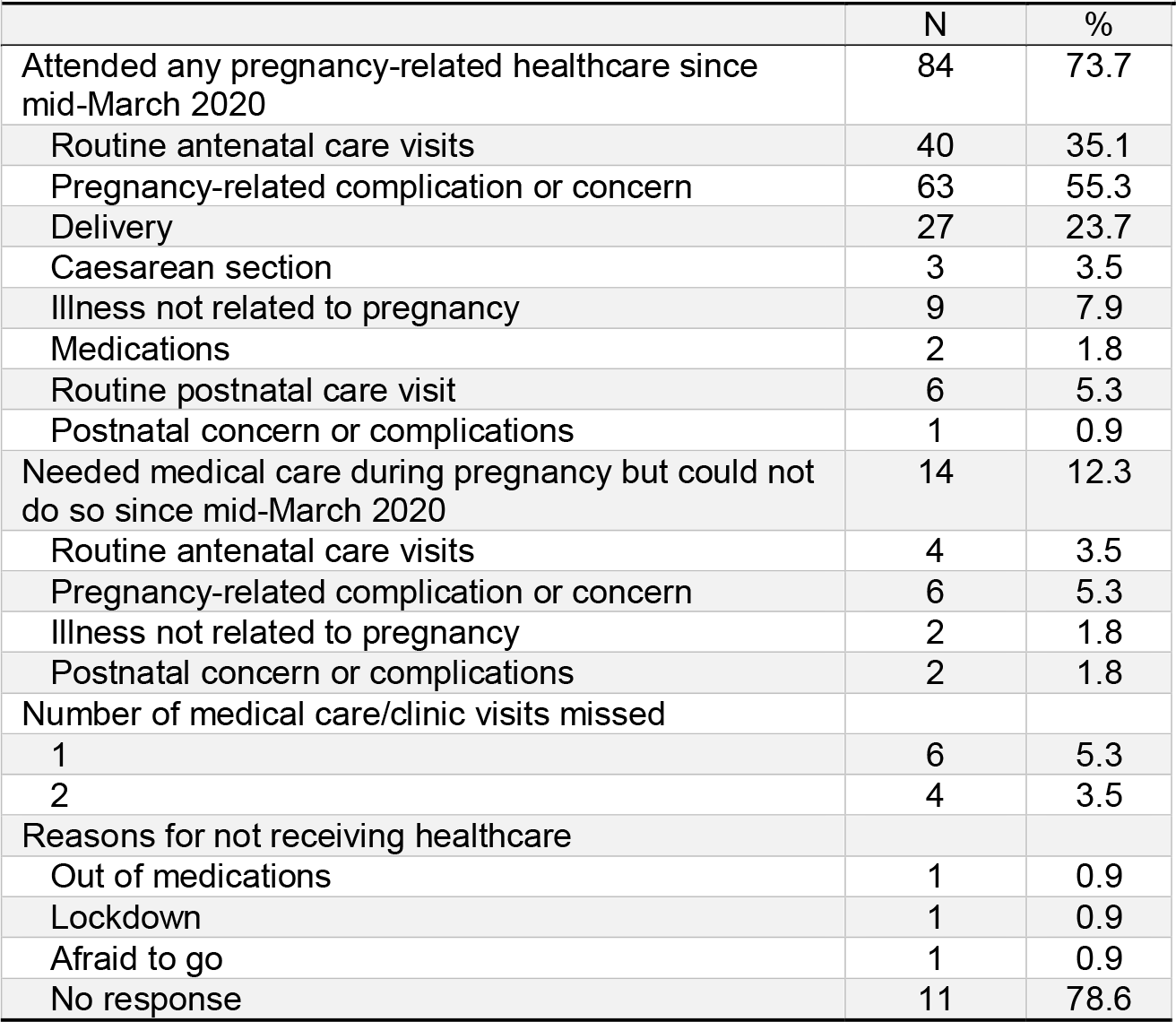
Pregnancy healthcare access, Ethiopia, August – September 2021 (N=114)

## Discussion

In this cross-sectional survey of community members in Eastern Ethiopia, nine of ten participants cited handwashing and facemask use as personal measures to prevent COVID-19, consistent with other studies conducted in Ethiopia.^12, 28, 36^ These measures have been shown in other settings to be associated with a substantial reduction in COVID-19 incidence.^37^ Conversely, wearing medical gloves was mentioned the least as a preventive measure for COVID-19. COVID-19 is primarily spread through exposure to respiratory droplets carrying infectious virus from coughs or sneezes, and transmission via contact with fomites (contaminated objects or surfaces) is possible, but the risk is generally considered to be low.^38^ The World Health Organization has warned that gloves use may have limited protective effectiveness for community members against SARS-CoV-2.^26, 39^ Regular use of gloves may provide a false sense of protection, and their incorrect use may favor SARS-CoV-2 transmission.^26^

Participants from rural areas were significantly more likely than those from urban areas to cite social distancing, avoiding crowded places, traveling, going out, and physical greetings to prevent COVID-19, which are among the most effective ways to prevent SARS-CoV-2 transmission.^37, 40^ These findings align with a study conducted in Harari, which demonstrated that individuals residing in rural areas were more likely to adhere to COVID-19 prevention measures, including handwashing, staying at home, maintaining social distancing, and wearing a facemask.^41^ These findings suggest that social distancing and reducing contact activities may be more feasible in rural areas, while participants from urban settings may encounter challenges in avoiding close contact with others.

Another possible explanation could be the disparity in access to healthcare information between rural and urban areas. Although our study did not assess specific sources of COVID-19 information, it was observed that television ownership was lower among participants from rural areas (19%) compared to those from urban households (87%). Moreover, community-based health insurance coverage was reported by a higher proportion of rural participants (59%) compared to urban participants (25%). Consequently, it is conceivable that rural participants acquired information on COVID-19 prevention from more reliable sources, such as community health workers or local health clinics. Prior studies in Ethiopia have reported television, social media, and radio as the most commonly cited sources of COVID-19 information, which were associated with higher levels of knowledge regarding COVID-19 transmission and prevention.^3, 12, 28^ Our study, however, did not find an association between television ownership and COVID-19 prevention knowledge. Although television is a common source of information, owning a television alone does not guarantee exposure to accurate and reliable COVID-19 information. Factors such as the frequency and quality of COVID-19 related programming, viewership habits, and media literacy could impact knowledge levels. In another study conducted in Harari Region using systematic random sampling, rural residents were found to be 1.62 (95% CI: 1.24, 2.10) times more likely to perceive a lower risk of COVID-19 compared to urban residents, potentially attributed to the lower population density in rural areas.^12^ Additional studies focusing on COVID-19 knowledge in Harari Region are needed to gain further insights into other factors influencing COVID-19 knowledge, including health infrastructure and resources, local context and cultural factors, and communication channels.

Knowledge of COVID-19 prevention was higher among participants aged 65 and older compared to younger participants, which is in accord with previous research.^29^ Older adults are at a greater risk of COVID-19-related complications, hospitalization, and mortality, particularly those with underlying health conditions.^42, 43^ According to data from the WHO, individuals aged ≥60 years accounted for over 80% of global COVID-19 mortality in 2020 and 2021.^44^ Other studies conducted in Ethiopia have also reported greater uptake of vaccination and other preventive measures among older adults compared to younger adults.^5, 28^ Although younger adults are less likely to be hospitalized with COVID-19 compared to older adults, some develop severe disease, and they can transmit the virus even when asymptomatic.^45^ Therefore, while the older population may already have better knowledge of COVID-19 prevention, interventions should also target younger age groups to address potential knowledge gaps and mitigate the risks associated with COVID-19 transmission. It is worth noting that Ethiopia has a young population, with a median age of 19.8 years, which is considerably lower than the median age of our study participants (40 years).^46^ Therefore, the higher knowledge levels observed among older adults in our study may be specific to this age group and may not necessarily reflect knowledge levels in younger age cohorts.

Households with higher monthly income had greater knowledge of COVID-19 prevention than households with the lowest income, which is consistent with previous studies in Ethiopia.^3, 28^ Eyeberu *et al*. found that individuals in Harari Region with higher income had greater perception of community risk of contracting COVID-19 than those in the lowest income group.^12^ However, among participants from rural areas, household size was found to be inversely associated with COVID-19 prevention knowledge, which is in contrast to a study of urban residents in northwest Ethiopia showing larger family sizes to be associated with greater COVID-19 knowledge and more positive attitudes towards COVID-19 prevention.^34^ In our study, none of the 40 participants from rural areas with household sizes of 9 or more members and only two of 134 (1.5%) participants from households of 7-8 members had completed secondary or college education. These proportions were lower than the 36.5% (19/52) of participants from households with only one or two individuals who had completed secondary or college education. Limited access to formal education in larger households could contribute to lower levels of COVID-19 knowledge.^47^

In April 2020, Ethiopia declared a state of emergency for COVID-19, which lasted for five months. The measures included suspending public gatherings such as religious congregations, sports, and concerts; quarantining travelers; requiring masking; closing schools and universities; and ordering some workers to work from home.^48, 49^ Additionally, other public health measures were implemented, such as restricting taxi and mass transit services, restricting long-distance travel to and from Addis Ababa, closing land borders, pardoning prisoners, postponing elections, and disseminating COVID-19 information via various media, including billboards and text messages.^48, 49^ Ethiopia also established exclusive facilities for COVID-19 healthcare and repurposed non-healthcare facilities as isolation centers and hospitals for COVID-19 care.^48^ In the Harari Region, we found that the most recognized community or government measures to prevent COVID-19 were closures of schools and universities and advice to avoid gatherings. More urban participants were also aware of government COVID-19 sensitization campaigns and advice to stay home, which may be attributed to exposure to multi-media messaging in cities.^28^

Our study also evaluated healthcare access for young children and pregnant women after the first COVID-19 case was reported in Ethiopia in March 2020. From mid-March to August-September 2021, over half of participants with children under five and three-quarters of pregnant women were able to attend healthcare visits, but one in ten missed at least one clinic visit due to concerns about contracting SARS-CoV-2. These findings align with a study in Southwest Ethiopia, which reported a significant reduction in family planning, antenatal care visits, healthcare facility births, and newborn vaccinations during the pandemic.^50^ During the pandemic in 2020 and 2021, there was a notable decline in childhood vaccination coverage, particularly in low and middle-income countries.^51^ Ethiopia also experienced disruptions in scheduled supplemental vaccination activities, including the postponement of a nationwide measles preventive vaccination campaign in April 2020.^52^ Another study in Ethiopia, demonstrated a significant decrease in childhood vaccinations, attributed to government lockdowns and inadequate supply by local providers and suppliers. To mitigate the impact of the pandemic on childhood vaccination coverage and ensure the continuity of essential healthcare services for children and pregnant women, it is recommended to facilitate the vaccination process by reducing waiting times at health centers, addressing parents’ concerns and fears related to COVID[19, improving vaccine availability, and promoting access in remote areas.^51^

This cross-sectional study has limitations, such as the lack of causal inference or temporality assessment, and potential social desirability bias in responses regarding COVID-19 prevention knowledge. As with any cross-sectional survey, the possibility of interviewer bias in conducting interviews cannot be completely ruled out. However, we took proactive measures to minimize this potential bias including comprehensive training provided to interviewers, rigorous recruitment processes for selecting qualified interviewers, and regular supervision of interviews by a data collection expert. Nonetheless, our study included a large sample of urban and rural community members, which provided the power to examine the data in several ways.

## Conclusion

In Harar and Kersa, Ethiopia, nine of ten community members knew that handwashing and mask-wearing could prevent COVID-19, but fewer identified avoiding crowded places and social distancing as prevention measures. Participants from rural areas demonstrated higher knowledge of COVID-19 prevention than those from urban areas. These findings suggest the importance of targeted outreach and community-engaged messaging to promote prevention measures, especially among younger, unemployed, and less educated individuals. In addition, promoting the safety and efficacy of COVID-19 vaccines through tailored communication strategies involving community leaders is necessary. Understanding these factors can aid authorities in developing effective education programs to control infectious disease transmission in households and communities.

## Data Availability

Data and questionnaire are publicly available at: https://doi.org/10.15139/S3/CZO1IX. Dheresa, Merga; Muir, Jonathan A.; Madewell, Zachary J.; Getachew, Tamirat; Daraje, Gamachis; Mengesha, Gezahegn; Whitney, Cynthia G.; Assega, Nega; Cunningham, Solveig A., 2023, "COVID-19 Impact Data for the CHAMPS HDSS Network: Data from Harar and Kersa, Ethiopia", https://doi.org/10.15139/S3/CZO1IX, UNC Dataverse, V1, UNF:6:lhiZWlzO4Cb2liqSV5jtSA== [fileUNF].

## Acknowledgements

We are grateful to the study participants who contributed their time in responding to our survey. We are also indebted to the fieldworkers in the data collection team that contacted household representatives and collected to data presented herein.

## Ethical approval

This study was conducted according to the guidelines in the Declaration of Helsinki; all procedures involving research study participants, including digital data collection using tablets that were programmed with the corresponding survey instruments, were approved by the Institutional Health Research Ethics Review Committee (IHRERC), College of Health and Medical Sciences, Harar Campus, Ethiopia; approval reference number Ref.No.IHRERC/127/2021. Written informed consent was obtained for participants who were able to read and write. For participants who were unable to read or write, the informed consent statement was read and oral informed consent from the participant was obtained, recorded, and witnessed. These procedures for obtaining written or oral informed consent were approved by the IHRERC, College of Health and Medical Sciences, Harar Campus, Ethiopia; approval reference number Ref.No.IHRERC/127/2021.

## Authors’ contributions

Conceptualization (MD, TG, GD, GM, NA), data curation (MD, TG, GD, GM, NA), formal analysis (ZM, JM), investigation (MD, TG, GD, GM, NA), methodology (MD, ZM, JM, TG, GD, GM, CW, NA, SC), project administration (MD, TG, GD, GM, CW, NA, SC), resources (MD, TG, GD, GM, NA), supervision (MD, CW, NA, SC), visualization (ZM, JM), writing – original draft preparation (ZM, JM), writing – review & editing (MD, ZM, JM, TG, GD, GM, CW, NA, SC).

## Availability of data and materials

Data and questionnaire are publicly available at: https://doi.org/10.15139/S3/CZO1IX.

Dheresa, Merga; Muir, Jonathan A.; Madewell, Zachary J.; Getachew, Tamirat; Daraje, Gamachis; Mengesha, Gezahegn; Whitney, Cynthia G.; Assega, Nega; Cunningham, Solveig A., 2023, “COVID-19 Impact Data for the CHAMPS HDSS Network: Data from Harar and Kersa, Ethiopia”, https://doi.org/10.15139/S3/CZO1IX, UNC Dataverse, V1, UNF:6:lhiZWlzO4Cb2liqSV5jtSA== [fileUNF].

## Informed consent

Written informed consent was obtained for participants who were able to read and write. For participants who were unable to read or write, the informed consent statement was read and oral informed consent from the participant was obtained, recorded, and witnessed.

## Disclaimer

The findings and conclusions in this report are those of the authors and do not necessarily represent the views of the US Centers for Disease Control and Prevention.

