## Supplement for "Knowledge of COVID-19 prevention in Eastern Ethiopia"

**Additional File 1**

| Table S1. Principal components factor analysis of knowledge of COVID-19 prevention variables, Ethiopia, August – September 2022 (N=880). | |
| --- | --- |
| Characteristic | Eigenvalue |
| Handwashing | 0.32 |
| Sanitizer use | 0.47 |
| Mask use | 0.16 |
| Avoid handshake/physical greeting | 0.67 |
| Avoid travel | 0.85 |
| Avoid going out | 0.85 |
| Avoid crowded places | 0.56 |
| Two-meter social distancing | 0.64 |
| *Eigenvalue* | 2.96 |
| Explained variance | 46% |

| Table S2. Generalized variance inflation factors of independent variables for analysis of associations between demographic characteristics and knowledge of COVID-19 prevention index, Ethiopia, August – September 2021 (N=880). | | | |
| --- | --- | --- | --- |
| Variable | **Overall** | **Urban (Harar)** | **Kersa (Rural)** |
| Age group | 1.202976 | 1.304589 | 1.197737 |
| Sex | 1.201439 | 1.16618 | 1.350462 |
| Residence | 1.696989 | -- | -- |
| Ethnicity | 1.363647 | 1.249853 | 1.50655 |
| Religion | 1.331012 | 1.319172 | 1.344809 |
| Marital status | 1.172729 | 1.249888 | 1.18851 |
| Education | 1.32604 | 1.216206 | 1.512683 |
| Occupation | 1.204472 | 1.118165 | 1.352122 |
| Health insurance | 1.192512 | 1.041689 | 1.247354 |
| Household size | 1.115762 | 1.089345 | 1.177772 |
| Children under 5 | 1.199537 | 1.233367 | 1.212692 |
| Adults over 60 | 1.126663 | 1.208579 | 1.11873 |
| Pregnant women | 1.098367 | 1.143196 | 1.131232 |
| Household member tested positive for COVID-19 | 1.075578 | 1.068938 | 1.212678 |
| Monthly income | 1.120503 | 1.107106 | 1.22044 |
| Has television | 1.687174 | 1.141773 | 1.713272 |

**Figure S1**. Knowledge of individual and community/government measures to prevent COVID-19, Harar and Kersa, Ethiopia, August – September 2021 (N=880). Error bars represent 95% confidence intervals.

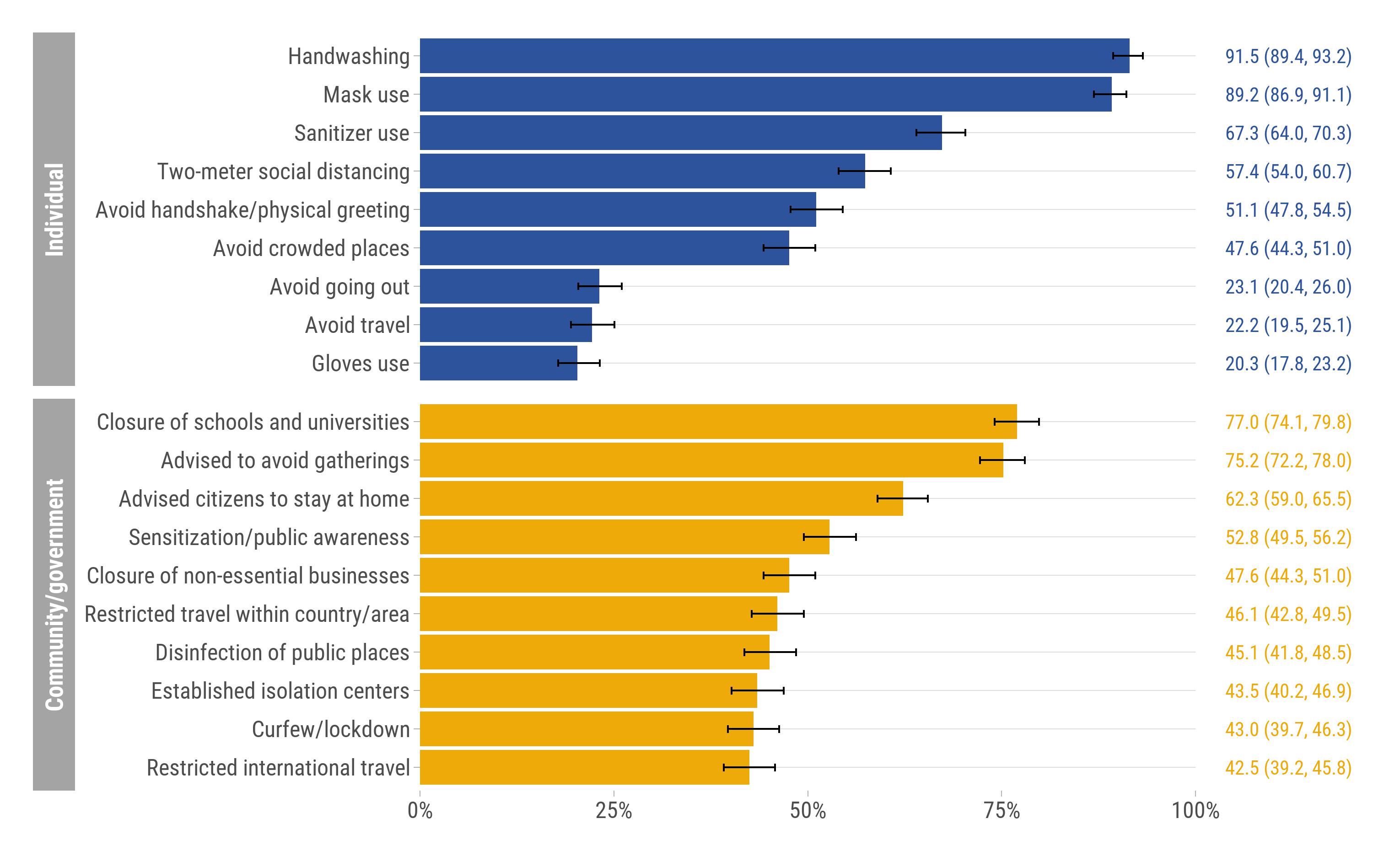

**Figure S2**. Unadjusted associations between demographic characteristics and knowledge of COVID-19 prevention index^a^ stratified by urban (Harar, N=440) and rural (Kersa, N=440) residence, Ethiopia, August – September 2021.

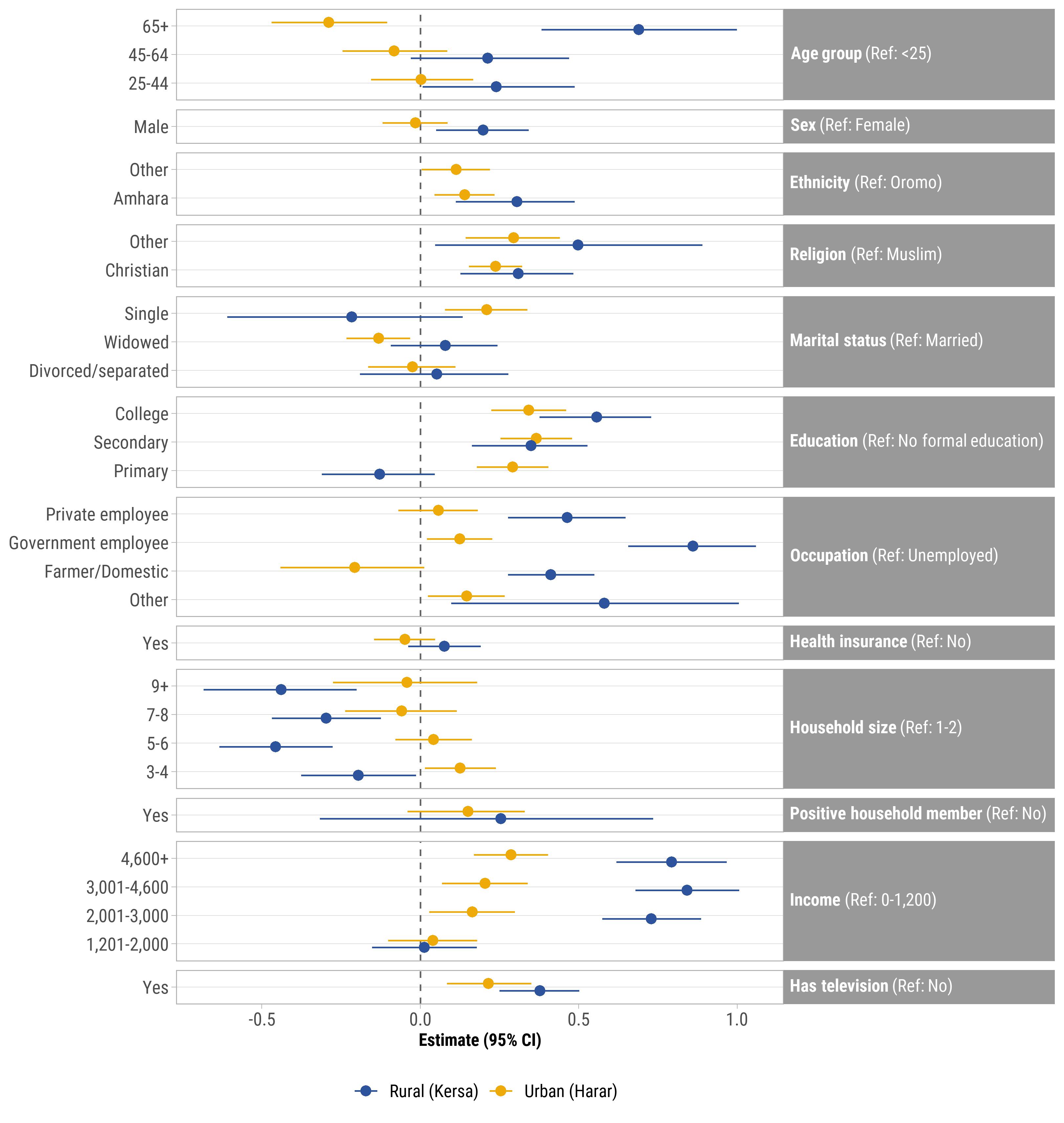

Points represent β coefficients and error bars represent 95% confidence intervals.
^a^ Knowledge of prevention was derived from principal components analysis and includes: handwashing, sanitizer use, mask wearing, avoiding handshaking/physical greeting, avoiding travel, avoiding going out, avoiding crowded places, and two-meter social distancing

**Figure S3**. Adjusted^a^ associations between respondent demographic characteristics and knowledge of COVID-19 prevention index^b^ stratified by urban (Harar, N=440) and rural (Kersa, N=440) residence, Ethiopia, August – September 2021.

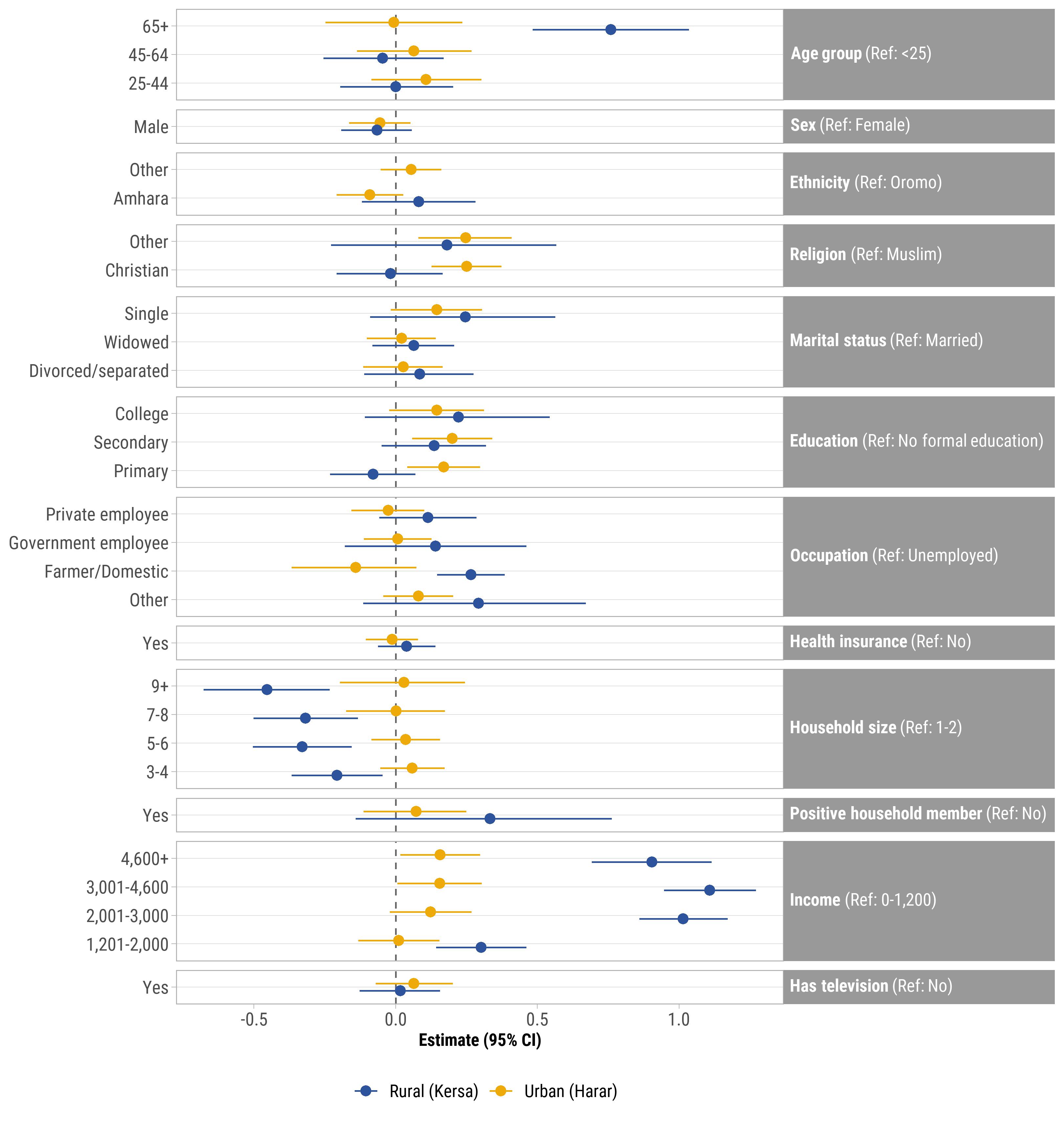

Points represent β coefficients and error bars represent 95% confidence intervals.
^a^ Adjusted for all other variables in the model.

^b^ Knowledge of prevention was derived from principal components analysis and includes: handwashing, sanitizer use, mask wearing, avoiding handshaking/physical greeting, avoiding travel, avoiding going out, avoiding crowded places, and two-meter social distancing
